# Remote virtual reality assessment elucidates self-blame-related action tendencies in depression

**DOI:** 10.1101/2022.03.17.22272498

**Authors:** Suqian Duan, Lucia Valmaggia, Diede Fennema, Jorge Moll, Roland Zahn

## Abstract

Darwin stated that humans have a strong and involuntary tendency to perform certain actions when a specific state of mind is induced. Such “action tendencies” are key to understanding the maladaptive impact of self-blame-related feelings in depression. For example, feeling like “hiding” and “creating a distance from oneself” in a text-based task were previously associated with recurrence risk in remitted depression. Despite their functional importance, action tendencies have not been systematically investigated in current depression, which was the aim of this pre-registered study. To this end, we developed the first virtual reality (VR) assessment of blame-related action tendencies and compared current depression (n=98) with control participants (n=40). The immersive VR-task, pre-programmed on devices sent to participants’ homes, used hypothetical social interactions, in which either participants (self-agency) or their friend (other-agency) were described to have acted inappropriately. Good concurrent validity was demonstrated for VR-versus the text-based task. As predicted, compared with controls, people with depression showed a maladaptive profile: Particularly in the other-agency condition, rather than feeling like verbally attacking their friend, they were prone to feeling like hiding, and punishing themselves. The depression group showed a more pronounced slowing of response times in the other-versus the self-agency condition, which may reflect a stronger ambivalence about externalising blame in depression. Interestingly, feeling like punishing oneself was associated with a history of self-harm but not suicide attempts. Current depression and self-harm history were thus linked with distinctive motivational signatures, paving the way for remote VR-based stratification and treatment.

**General Scientific Summary:** As precursors of social actions, action tendencies, such as feeling like hiding when experiencing shame or guilt, provide the key link between depressive emotions and behaviour. Here, we developed a novel virtual reality task and used it in a large sample of people with depression to unveil a distinctive pattern of such action tendencies. As predicted, we found a maladaptive pattern of action tendencies in people with depression that were specifically associated with self-harm. Our findings thereby elucidate a so far unexplored key aspect of psychopathology in depression and provide important targets for the development of novel future psychological interventions tackling self-harm-related motivations and depression.

## Introduction

Proneness to overgeneralised self-blaming emotions plays a central role in cognitive models of depression (Abramson et al., 1978; Janoff-Bulman, 1979). Self-blaming emotions are associated with a tendency to perform specific actions such as hiding and apologising (Duan et al., 2022). In his seminal chapter “The Expression of the Emotions in Man and Animals”, Darwin stated that humans have a strong and involuntary tendency to perform certain actions when a specific state of mind is induced and emphasised the evolutionary importance of these tendencies (Darwin, 1872). Such implicit tendencies, so called “action tendencies”, describe a cognitive and motivational state before an action is taken (Haidt, 2003), thereby providing the crucial link between emotion and action. Due to its immersive nature, virtual reality (VR) is ideally suited to uncover usually implicit experiences. Here, we employed a novel remote VR task to investigate the role of blame-related action tendencies in depression. Identifying depression-related maladaptive action tendencies is an essential step towards understanding the link between emotions and actions and developing novel interventions and cognitive markers.

Over the past 50 years, a growing body of research has focused on the importance of self-blaming emotions in the development of depression (e.g. Power & Dalgleish, 2015). More specifically, the feeling of overgeneralised guilt (O’Connor et al., 2002) and a bias towards self-contempt have been found in people with depression even on remission of their symptoms (Zahn et al., 2015). These findings suggested self-blaming emotions as vulnerability factors for depression. However, it might be misleading to understand the relationship between self-blaming emotions and depression based on the emotion labels alone. For example, there are different forms of guilt which can be either adaptive or maladaptive. Adaptive guilt was associated with good social adjustment and was not related to depression vulnerability (Tangney et al., 2007). In contrast, an overgeneralised form of guilt was maladaptive and increased in major depressive disorder (O’Connor et al., 2002). Consequently, to understand self-blaming emotions in depression, one needs to unveil their link with motivating adaptive and maladaptive social actions.

These implied adaptive (e.g. “feeling like apologising”) and maladaptive social actions (e.g. “feeling like hiding”) have been referred to as “action tendencies” in the social psychology literature (Haidt, 2003). Maladaptive action tendencies have been defined as attempts to deny, hide or escape the emotion-inducing situation, whereas adaptive action tendencies are defined as implying reparative actions that help people face and deal with a difficult situation (Haidt, 2003; Tangney et al., 2007). It was shown that people with higher depressive symptoms exhibited higher levels of maladaptive action tendencies such as withdrawal compared with those with lower symptoms (Mu & Berenbaum, 2019). In support of this, our recent study also found that people with remitted depression had more maladaptive action tendencies including feeling like “hiding”, “creating a distance from oneself” and “attacking oneself” than those without a history of depression (Duan et al., 2022). These maladaptive action tendencies were related to either escaping the situation, denial of one’s identity, or self-punishment which might further contribute to depressogenic cognitive styles in stressful situations and ultimately increase the likelihood of developing depression. Indeed, maladaptive action tendencies were a prospective risk factor for recurrence risk in remitted major depressive disorder (Lawrence et al., 2021).

While maladaptive blame-related action tendencies were associated with vulnerability to depression (Lawrence et al., 2021), so far, their role in current depression and associated maladaptive behaviours is elusive. Specifically, self-harming and suicidal behaviours are of the highest clinical relevance in depression. Both have been associated with self-blaming emotions (Sheehy et al., 2019) and punishment-related behaviours (Stanicke, 2021), but the role of action tendencies in self-harming behaviours is unknown. In addition, previous measures of action tendencies used abstract verbal descriptions (Lawrence et al., 2021), which heavily relied on how well participants could imagine and contextualise these action tendencies. Also, these measures did not allow participants to act out their action tendencies, which further limited their immersiveness and engagement in the scenarios. VR-based assessment is a new experimental paradigm for psychometric evaluation compared with traditional paper-and-pencil or computerised tasks. VR scenarios were suggested to be safe and promising tools for cognitive assessments in people with psychiatric disorders (Falconer et al., 2016; Henry et al., 2012). The interactive and immersive nature of the VR environment renders it well suited in depicting blame-related social scenarios and allows to measure action tendencies in controlled experimental conditions.

For the present study, we developed such a novel VR task to assess blame-related action tendencies with the following aims: firstly, to validate the VR task as a self-assessment measure of blame-related action tendencies and self-blaming biases in people with current depression by comparing it against a shortened, optimised version of our previously reported text-based version (Duan et al., 2022); secondly, to probe our pre-registered hypothesis that people with current depression exhibit higher levels of maladaptive blame-related action tendencies, such as feeling like hiding and creating a distance from oneself (clinicaltrials.gov: NCT04593537, research question a.); and thirdly to investigate the hypothesis, that maladaptive action tendencies, specifically feeling like punishing oneself, are associated with a higher risk of self-harm and suicide attempt based on self-reported history.

## Methods

### Participants

This study was approved by the King’s College London PNM Research Ethics Subcommittee (Project Reference:HR-19/20-17589) and pre-registered NCT04593537 with a cross-sectional part which we reported in the current paper and a prospective prognostic study which will reported separately. All participants were compensated with a £25 Amazon voucher on completing the study or £15 for only completing the online baseline assessment. Participants were recruited via online advertising on social media as well as the King’s College London department circular. The eligibility of participants was determined by an online pre-screening questionnaire, which participants accessed via the online adverts. General inclusion criteria were: age of 18 years or over and being able to complete self-report scales orally or in writing. General exclusion criteria were: a personal or family history of schizophrenia, schizoaffective disorder, or bipolar disorder, a personal history of psychotic symptoms, drug or alcohol abuse over the last 6 months, a suspected central neurological condition, a planned or current pregnancy, or currently being treated by a mental health specialist in secondary care. In addition, participants were excluded if they had hypomanic symptoms [The Hypomanic Checklist-16 (Forty et al., 2010) score > 8, symptoms lasting ≥ 2 days] and endorsed two of the first three screening questions of the Composite International Diagnostic Interview bipolar screening scale (Kessler et al., 2006). Supplementary Figure 1 shows exclusions reasons for participants and a flow chart of the participant recruitment. Specific depression group inclusion criteria were: at least a moderately severe major depressive syndrome at screening [The Patient Health Questionnaire-9 (PHQ-9, Kroenke et al., 2001) score ≥ 15], and early treatment resistance to antidepressants [here defined as having tried at least one antidepressant medication in primary care according to (Fekadu et al., 2018)]. Specific control group exclusion criteria were: a personal or family history of depression, a personal history of taking antidepressants, and a PHQ-9 score>9. Specific control group inclusion criteria were: matching for demographic variables with the depression group.

In total, 897 participants completed the online pre-screening questionnaires, 164 participants were eligible for the depression and 198 for the control group. One hundred-and-one participants with depression and 40 control participants agreed to participate and completed the online baseline assessment of the study; with 98 in the depression group and all control participants completing the VR task.

### Statistical power

The sample size of the depression group was primarily powered for the prospective prognosis study, which will be reported separately (clinicaltials.gov: NCT04593537). We aimed for a control group sample size of at least 35 to allow for reliable standard deviation estimates which are needed for effect size estimates in feasibility studies in the absence of known effect sizes (Teare et al., 2014). For our main question in this paper (aim 2), our overall attained sample size of n=138 allowed us to detect a Cohen’s F2 effect size of ≥0.16 (i.e. medium effect size), corresponding to η2≥.14 with 90% power at a 2-sided p≤.05 in the MANOVA with 10 action tendency outcome variables and one predictor variable (group, df=[127,10], G-Power software version 3.1.9.7).

### Assessment of clinical characteristics

In the online baseline assessment, questionnaires were developed to collect participants’ demographic information as well as to measure their depressive and anxiety symptoms [the Quick Inventory of Depressive Symptomatology – self-reported -16 [QIDS-SR-16, (Rush et al., 2003)], Maudsley-Modified Patient Health Questionnaire [MM-PHQ-9, (Harrison et al., 2021)] and Generalised Anxiety Disorder-7 Scale [GAD-7, (Spitzer et al., 2006)]. Participants in the depression group were also asked to select the antidepressants they were currently taking or had taken in the past two months as well as the antidepressants they had taken prior to that. For each antidepressant participants had chosen, they were required to specify the dosage, duration, effectiveness, side effect(s) and the reason why they have stopped taking this antidepressant (if applicable). In addition, at baseline participants had to report the number of previous episodes, age at onset, the duration of their most recent depressive episode, a history of self-harm and of suicide attempts as well as to report specific symptoms.

### Virtual reality assessment of blame-related action tendencies

The virtual reality environment was developed by the King’s College London, Institute of Psychiatry, Psychology and Neuroscience VR Research Lab in Unity, deployed to Oculus Go. The design of the VR task was based on the value-related moral sentiment task (VMST) which has been described in (Zahn et al., 2015). Scenarios used in the VR task were chosen from a previous study in which participants were interviewed and gave examples of situations when they experienced blame-related emotions (Green et al., 2013). Scenarios were only selected if they were judged to trigger blame-related emotions across individual and cultural contexts. In order to make the task both brief and sensitive to detect blame-related emotions, 15 scenarios were chosen, with each scenario being displayed in both the self-agency and other-agency condition (30 trials in total, 15 per condition). In the self-agency condition, the participant acted counter to social and moral values in the interaction with their friend (the participant was the agent). In the other-agency condition, the participant’s friend was the agent and the participant the recipient of the action.

At the beginning of the VR task, participants were presented with the welcome message and asked to enter their participant ID. As the task began, participants were taken to a scene in the street, a shopping centre, or a coffee shop, while a narrator described a hypothetical social scenario that happened between the participants and their friends. The full list of the narrative of the hypothetical social scenarios is included in the supplementary Table 1. One example of the narrative in the self-agency condition is “You drove your friend’s car, caused an accident and damaged it”. Participants then saw the VR avatar entering the scene and moving towards them. The narrator said, “You just saw your friend, what would you feel like doing?”. Participants saw choice options displayed on the screen and asked to choose one of them (**Figure 1**). Among the choices were seven different action tendencies: “Verbally attacking my friend”, “Punishing myself”, “Apologising”, “Hiding”, “Creating distance from my friend”, “Creating distance from myself”, and “Other/no action”. As soon as participants made their choices using their virtual hands, the display changed accordingly to perform the corresponding actions. For “Feeling like verbally attacking my friend”, participants were asked by the narrator to read out aloud a sentence: “How could you do this to me?” with a raised voice to the friend. For “Feeling like punishing myself”, participants were taken to a cartoon depiction of someone who was irritated with himself. Participants who chose “Feeling like apologising” were asked by the narrator to say out aloud a sentence: “I am sorry, what can I do to make this up to you?”. Participants who chose “Feeling like hiding” were taken to a visual display which made them feel they were hiding either behind a wall or under a table. If participants’ choice was “Feeling like creating a distance from your friend”, the visual display changed in that the friend’s avatar was depicted from a distance. For “feeling like creating a distance from yourself”, the visual display moved to a bird’s eye perspective with a depiction of oneself being seen from above together with the friend. For “Other action/no action”, the screen went black for five seconds and then went to the next trial. At the end of each trial, participants were asked to rate their levels of self-blame (how strongly would you blame yourself?) and other-blame (how strongly would you blame your friend?) from 1 to 7, where 1 corresponded to “not at all” and 7 to “very much”, shown on the screen as visual analogue scroll bars. Depending on the speed of participants completing the task, the VR task took around 25 minutes in total. Response time taken for participants to choose different action tendencies was recorded by measuring the duration between action tendency options displayed on the screen and participants selecting one of the action tendencies. Screenshots and video of the VR task can be found in Figure 1 and https://youtu.be/agWahwvYDXc.

**Figure 1.**
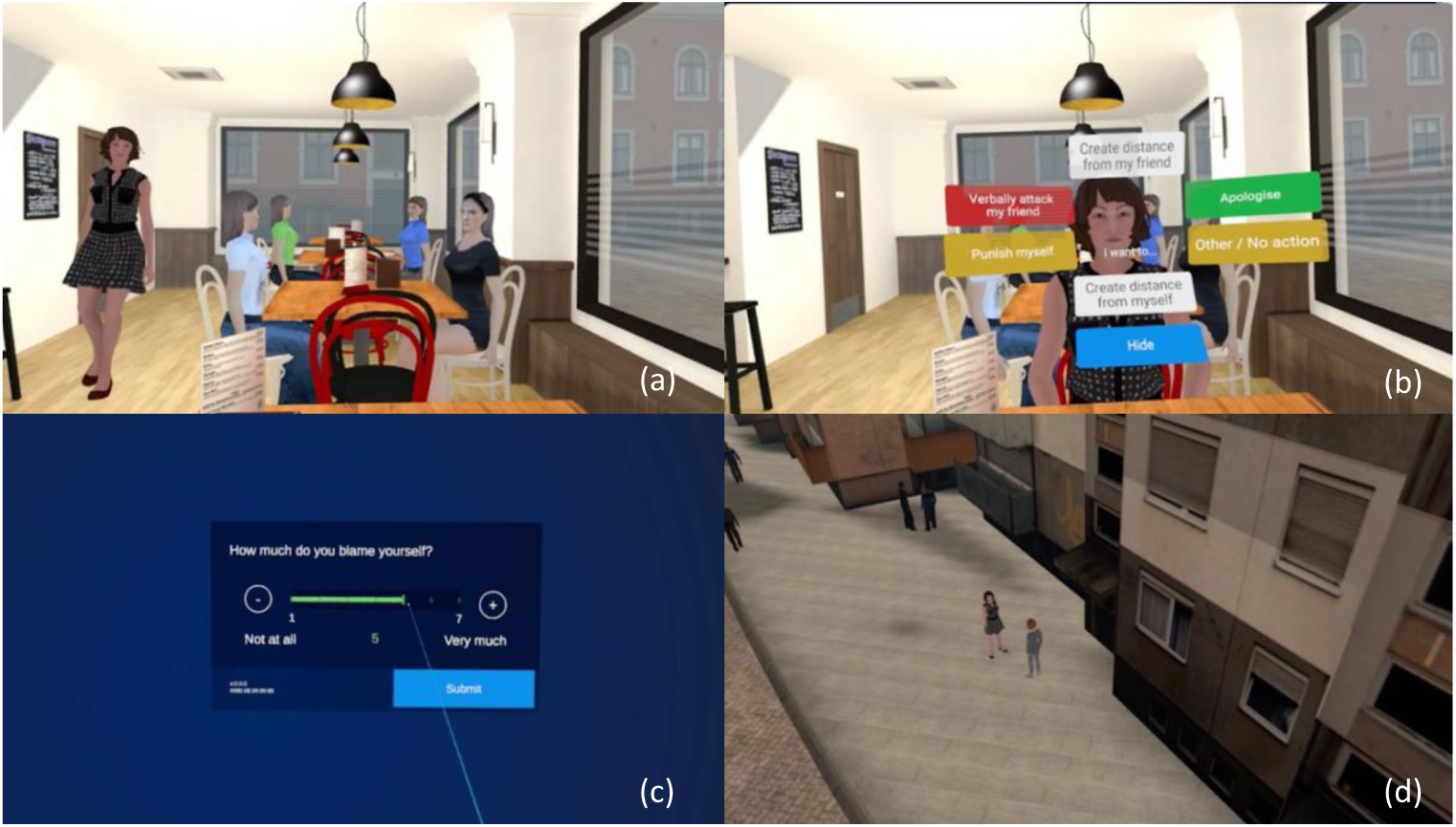
Screenshots of the VR task Panel (a) shows that participants meet their friends in a coffee shop, panel (b) depicts the action tendency buttons that participants were asked to choose from using their virtual hands. Panel (c) shows the screen that participants saw when for their blame ratings. Panel (d) depicts how participants viewed themselves and their friend from a bird-eye perspective after they chose creating a distance from themselves.

### Text-based assessment of blame-related action tendencies

The text-based action tendency task was completed by participants to compare with the virtual reality action tendency task, either using Excel Macro or using PsychoPy (Peirce et al., 2019). This task has been validated in our previous study (Duan et al., 2022) but here, we used a modified, shortened version with 54 trials (27 trials in the self-agency condition and 27 trials in the other-agency condition. The original task consisted of 180 statements, resulting in a long session duration, and not all social concepts were deemed relevant to overgeneralised feelings of self-blame. In addition to shortening the task, the statements themselves were optimised by excluding negated positive social concepts (e.g. “does not act generously”) and replacing “does act [social concept]” with “is [social concept]).

In the task, participants were presented with hypothetical social interactions between them and their friend, during which either the participant (self-agency condition) or the participant’s friend (other-agency condition) acted counter to social and moral values. To personalise the statements, participants were asked to name their friend in the initial set-up. For each interaction/trial, participants were asked to choose one emotion from six moral emotions: shame, guilt, contempt/disgust towards oneself, contempt/disgust towards friend, indignation/anger towards friend, or no/other feeling, and one action tendency from six action tendencies: apologising, hiding, creating a distance from oneself, creating a distance from friend, verbally or physically attacking friend, or no/other action. In addition, participants were asked to indicate how strongly they would blame themselves and their friend for the imagined behaviour on a scroll bar from 1 (“not at all”) to 7 (“very much”).

### Procedure

This study was conducted fully remotely between June 2020 to June 2021, thereby allowing us to conduct the study during the COVID-19 pandemic. After participants enrolled in the study, they received the links to complete the online baseline assessment, the text-based action tendency task by email and a VR headset by courier. Participants were asked to complete the online baseline assessment first, followed by the text-based action tendency task and finally the VR action tendency task following the instructions provided by the researcher. The VR task was completed by participants unsupervised, however, if participants had any questions during the completion of the task, the researcher was available on the phone or via video conference if needed. Participants sent the VR headset back to the researcher after they completed all the tasks.

### Data analysis

All data were analysed using IBM SPSS statistics version 27. Means and standard deviations were calculated for the proportion of choosing each action tendency for each participant in each condition (self-agency and other-agency). The effect of depressive symptoms (QIDS-SR-16 score), anxiety symptoms (GAD-7 score) and medication status on average action tendency proportions in the depression group were examined by a multivariate analysis of variance (MANOVA). For aim 1, the newly developed VR task was validated by comparing it with the text-based action tendency task. Self-blame rating bias measures were calculated for each participant and agency-condition (self-vs. other-) by subtracting the average other-blame ratings from self-blame ratings. Pearson correlation analyses were carried out to examine the relationship between pairs of corresponding action tendencies and self-blaming bias measures in the text-based and VR-based tasks. The reference correlation was computed by taking the average of the Pearson correlation coefficients across all action tendencies, except the action tendency of interest. The correlation for each action tendency was compared against the reference correlation using the Fisher-Z-transformation. A significant Pearson correlation and Fisher’s z value indicated construct validity of the novel VR task when compared with the text-based task.

The VR task was further validated by investigating the relationship between action tendencies and self-blame rating bias measures using a MANOVA, with different action tendencies as dependent variables and self-blame rating bias measures in both conditions as independent variables. For aim 2, the effects of group on action tendencies were examined using a MANOVA with different action tendencies as dependent variables and group as the only independent variable. For aim 3, the role of punishing oneself in relation to past self-harm and suicide attempt were examined by another MANOVA with punishing oneself in both conditions as dependent variables and self-harm and suicide attempt as the independent variables. For all MANOVAs, post-hoc univariate tests were carried out if significant multivariate effects were found. Multiple comparison correction at a two-sided p=.05 using the Benjamini–Hochberg procedure was employed for all post-hoc univariate tests. Repeated measure analysis of variance (ANOVA) was used to examine the differences of response time taken to choose action tendencies in each agency condition and group.

## Results

### Clinical characterisation of the participants

The characteristics of participants with depression and control participants are shown in Table 1. There were no significant group differences on any demographic variables including age, years of education, sex ratio, ethnicity, native language, or employment status. The clinical characteristics of participants with depression are shown in Table 2. Most of them had one to two treatment failures as defined by the Maudsley Staging Method (Fekadu et al., 2018), a duration of their current depressive episode of ≤12 months, depressive symptoms that fell into the severe range, and were currently taking a single SSRI as their antidepressant medication.

**Table 1.**
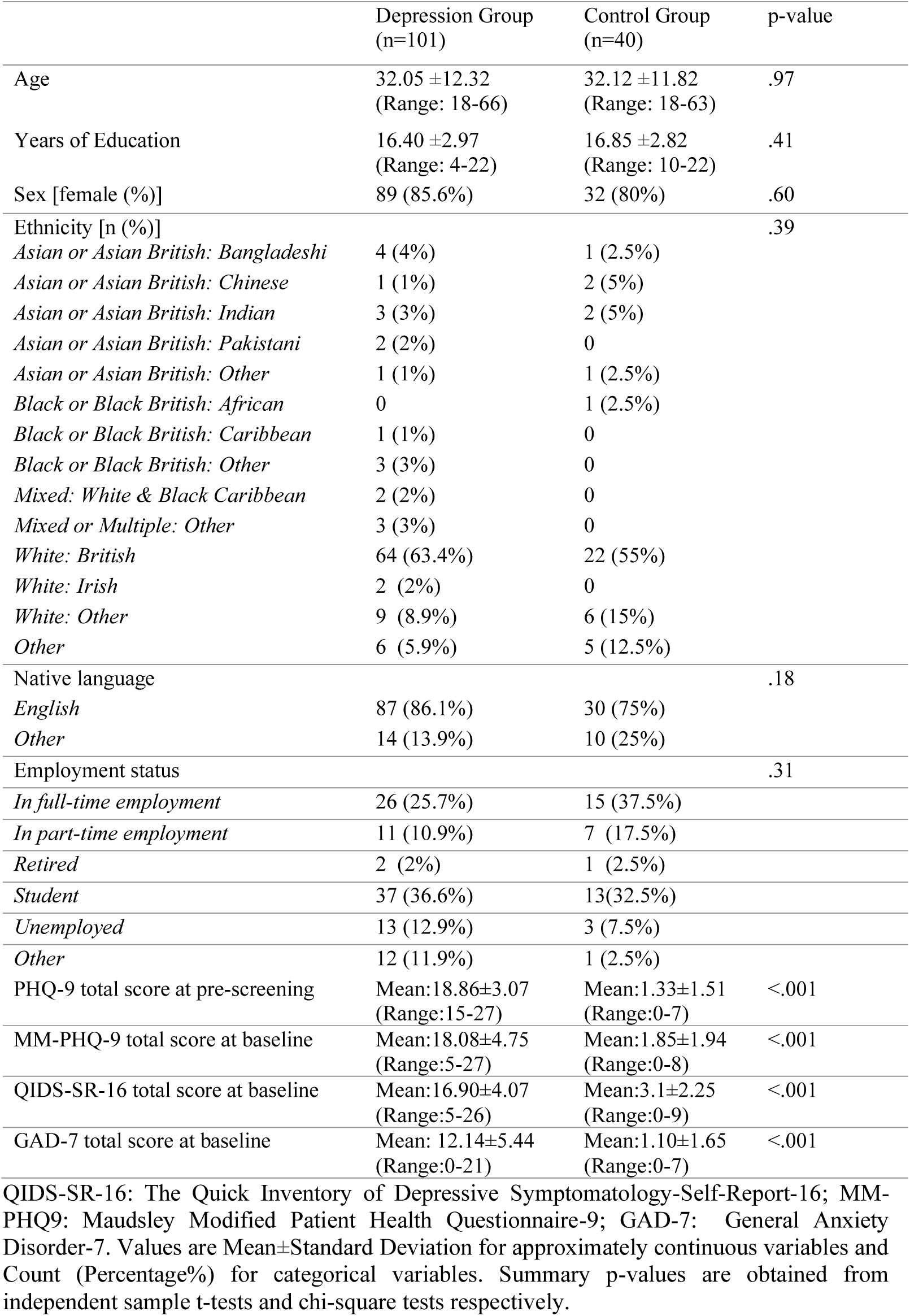
Comparison of participants’ demographic characteristics and clinical characteristics

|  | Depression Group<br>(n=101) | Control Group<br>(n=40) | p-value |
| --- | --- | --- | --- |
| Age | 32.05 ±12.32<br>(Range: 18-66) | 32.12 ±11.82<br>(Range: 18-63) | .97 |
| Years of Education | 16.40 ±2.97<br>(Range: 4-22) | 16.85 ±2.82<br>(Range: 10-22) | .41 |
| Sex [female (%)] | 89 (85.6%) | 32 (80%) | .60 |
| Ethnicity [n (%)] |  |  | .39 |
| *Asian or Asian British: Bangladeshi* | 4 (4%) | 1 (2.5%) |  |
| *Asian or Asian British: Chinese* | 1 (1%) | 2 (5%) |  |
| *Asian or Asian British: Indian* | 3 (3%) | 2 (5%) |  |
| *Asian or Asian British: Pakistani* | 2 (2%) | 0 |  |
| *Asian or Asian British: Other* | 1 (1%) | 1 (2.5%) |  |
| *Black or Black British: African* | 0 | 1 (2.5%) |  |
| *Black or Black British: Caribbean* | 1 (1%) | 0 |  |
| *Black or Black British: Other* | 3 (3%) | 0 |  |
| *Mixed: White & Black Caribbean* | 2 (2%) | 0 |  |
| *Mixed or Multiple: Other* | 3 (3%) | 0 |  |
| *White: British* | 64 (63.4%) | 22 (55%) |  |
| *White: Irish* | 2 (2%) | 0 |  |
| *White: Other* | 9 (8.9%) | 6 (15%) |  |
| *Other* | 6 (5.9%) | 5 (12.5%) |  |
| Native language |  |  | .18 |
| *English* | 87 (86.1%) | 30 (75%) |  |
| *Other* | 14 (13.9%) | 10 (25%) |  |
| Employment status |  |  | .31 |
| *In full-time employment* | 26 (25.7%) | 15 (37.5%) |  |
| *In part-time employment* | 11 (10.9%) | 7 (17.5%) |  |
| *Retired* | 2 (2%) | 1 (2.5%) |  |
| *Student* | 37 (36.6%) | 13 (32.5%) |  |
| *Unemployed* | 13 (12.9%) | 3 (7.5%) |  |
| *Other* | 12 (11.9%) | 1 (2.5%) |  |
| PHQ-9 total score at pre-screening | Mean:18.86±3.07<br>(Range:15-27) | Mean:1.33±1.51<br>(Range:0-7) | <.001 |
| MM-PHQ-9 total score at baseline | Mean:18.08±4.75<br>(Range:5-27) | Mean:1.85±1.94<br>(Range:0-8) | <.001 |
| QIDS-SR-16 total score at baseline | Mean:16.90±4.07<br>(Range:5-26) | Mean:3.1±2.25<br>(Range:0-9) | <.001 |
| GAD-7 total score at baseline | Mean: 12.14±5.44<br>(Range:0-21) | Mean:1.10±1.65<br>(Range:0-7) | <.001 |

QIDS-SR-16: The Quick Inventory of Depressive Symptomatology-Self-Report-16; MM-PHQ9: Maudsley Modified Patient Health Questionnaire-9; GAD-7: General Anxiety Disorder-7. Values are Mean ± Standard Deviation for approximately continuous variables and Count (Percentage%) for categorical variables. Summary p-values are obtained from independent sample t-tests and chi-square tests respectively.

**Table 2.** Clinical characteristics of depression group at baseline

|  |  |
| --- | --- |
| Number of failed treatments |  |
| 1-2 | 61 (60.4%) |
| 3-4 | 34 (33.7%) |
| 5-6 | 5 (5%) |
| 7-10 | 1 (1%) |
| Duration of current depressive episode |  |
| ≤12 months | 76 (75.2%) |
| 13-24 months | 10 (9.9%) |
| >24 months | 15 (14.9%) |
| Age at first onset | Mean±SD: 16.41±6.48<br>(Range:4-55) |
| Maudsley Staging Model<br>total score | Mean: 6.47±1.40<br>(Range:4-11) |
| Maudsley Staging Model<br>severity |  |
| Mild | 60 (57.7%) |
| Moderate | 40 (38.5%) |
| Severe | 1 (1%) |
| Current medication |  |
| Single SSRI | 58 (55.8%) |
| Single SNRI | 13 (12.5%) |
| Other | 14 (13.4%) |
| None | 19 (18.3%) |
| Self-reported co-morbid psychiatric conditions |  |
| PTSD | 10 (10.0%) |
| Anxiety disorders | 7 (6.9%) |
| Eating disorders | 7 (6.9%) |
| Personality disorders | 5 (4.9%) |
| OCD | 5 (4.9%) |
| ASD | 3 (3.0%) |
| ADHD | 1 (1.0%) |
| Other | 4 (4.0%) |

n=101. SSRI: Selective serotonin reuptake inhibitors; SNRI: Serotonin and norepinephrine reuptake inhibitors. PTSD: posttraumatic stress disorder; OCD: obsessive-compulsive disorder; ASD: autism spectrum disorder; ADHD: attention deficit hyperactivity disorder; Values are Mean ± Standard Deviation for approximately continuous variables and Count (Percentage%) for categorical variable.

### Descriptive statistics of action tendencies in the VR task

Means, standard deviations and standard errors for proportions of selecting different action tendencies are presented in Figure 2. Feeling like apologising was the most frequently chosen action tendency in the self-agency condition for both groups (0.58 and 0.65 for the depression and the control group). In contrast, feeling like creating a distance from one’s friend (0.30 for both groups) and other/no action (0.31 and 0.30 for the depression and the control group) were most commonly chosen in the other-agency condition.

**Figure 2.**
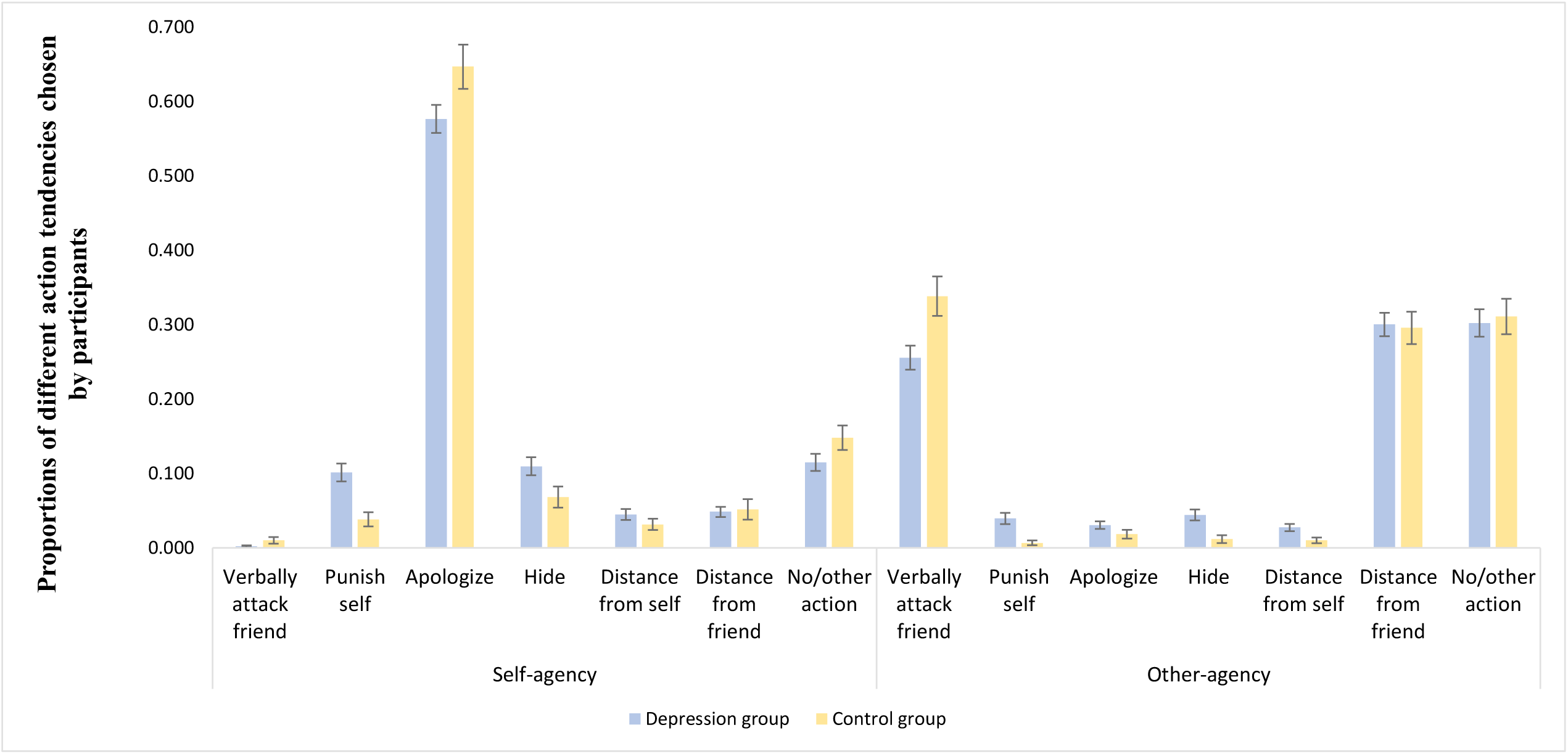
Proportions of different action tendencies chosen by participants N=138 (depression group: 98; control group: 40); error bars show standard errors of the proportions.

### Associations of VR action tendencies and clinical characteristics in the depression group

There was no multivariate effect of depressive symptoms (F(10,82)=.67, p=.75, Wilk’s Λ = .08, partial η2 = .08), anxiety symptoms (F(10,82)=.56, p=.84, Wilk’s Λ = .06, partial η2 = .06), nor medication status on action tendencies in the depression group (F(10,82)=.86, p=.57, Wilk’s Λ = .10, partial η2 = .10). This independence of action tendencies of symptom severity was further confirmed in exploratory univariate correlations: no significant correlation between any action tendency with either depressive or anxiety symptom levels was found in the depression group (see Supplementary Table 2).

### Validation of the VR task

Action tendencies and self-blame rating biases were compared between the VR- and text-based tasks to probe their construct validity (see Table 3). Overall, most action tendencies in the VR task were significantly correlated with those in the text-based task (r ranging between 0.27-0.55), except hiding in the self-agency condition and distancing from oneself in the other-agency condition. After the Fisher-Z-transformation, there were also significant differences found between these action tendencies and the reference correlations, showing that associations were specific for corresponding action tendencies. In addition, self-blame rating biases in the VR task were highly correlated with self-blame rating biases in the text-based task in both conditions (r=0.68 in the self-agency and r=0.75 in the other-agency).

**Table 3.** Pearson correlations between action tendencies and self-blaming biases in the text-based and VR action tendency task

|  | Self-agency |  |  |  | Other-agency |  |  |  |  |  |
| --- | --- | --- | --- | --- | --- | --- | --- | --- | --- | --- |
|  | Distancing<br>from oneself | Hiding | Apologising | Self-blame<br>rating biases | Distancing<br>from oneself | Hiding | Apologising | Attacking<br>friend | Distancing<br>from friend | Self-blame<br>rating biases |
| Pearson correlation | .43 | .17 | .42 | .68 | -.03 | .55 | .19 | .34 | .27 | .75 |
| p-value for Pearson<br>correlation | <.001*** | .07 | <.001*** | <.001*** | .77 | <.001*** | .03* | <.001*** | .003** | <.001*** |
| Fisher's z against reference<br>correlation | 4.84 | 1.29 | 5.57 |  | -0.35 | 6.20 | 1.85 | 4.41 | 3.13 |  |
| p-value for Fisher's z | <.001*** | .10 | <.001*** |  | 0.36 | <.001*** | 0.03* | <.001*** | <0.001** |  |

Fisher's z was computed against a reference correlation for each action tendency. The reference correlation was calculated by taking the average of the Pearson correlations between all action tendencies, except the action tendency of interest in the VR action tendency task and the action tendency of interest in the text-based action tendency task. For example, the reference correlation for SA distancing from oneself was calculated by averaging the correlations between all action tendencies except SA distancing from oneself (SA hiding, SA apologising, OA verbally attacking friend, OA apologising, OA hiding, OA distancing from oneself, OA distancing from friend) in the VR action tendency task and SA distancing from oneself in the text-based action tendency task. There was one reference correlation calculated for each action tendency. Self-blame rating bias was calculated by subtracting each participant's average other-blaming ratings from their average self-blaming ratings for each agency.
\*\*\*<.001; \*\*<.01, \*<.05 at a significance level of alpha=.05. n=121 in all analyses.

The VR task was further validated by demonstrating a significant association between multivariate action tendency profiles as outcomes and self-blaming biases as predictors across groups (self-agency condition: F(10,126)=7.19, p<.001, Wilk’s Λ = .36, partial η2 = .36; as well as other-agency condition: F(10,126)=10.45, p<.001, Wilk’s Λ = .45, partial η2 = .45). After applying multiple comparison correction, post-hoc univariate ANOVAs showed significant associations of feeling like punishing oneself and apologising with higher self-blaming biases in the corresponding self- and other-agency conditions. Furthermore, the association of agency-incongruent self-blaming biases (blaming oneself when one’s friend has acted badly) with a lower tendency to verbally attack one’s friend in the other-agency condition survived multiple comparison correction (see Table 4).

**Table 4.** The effects of self-blame rating biases on different action tendencies

| Dependent Variable |  | B | Std. Error | t-value | p-value | 95% Confidence Interval |  | Partial Eta Squared |
| --- | --- | --- | --- | --- | --- | --- | --- | --- |
|  |  |  |  |  |  | Lower Bound | Upper Bound |  |
| SA punish self | SA self-blame rating bias | 0.02 | 0.01 | 3.20 | .002***† | 0.01 | 0.04 | 0.07 |
|  | OA self-blame rating bias | 0.01 | 0.01 | 2.80 | 0.01** | 0.00 | 0.02 | 0.06 |
| SA apologise | SA self-blame rating bias | 0.04 | 0.01 | 3.41 | <.001***† | 0.02 | 0.07 | 0.08 |
|  | OA self-blame rating bias | -0.01 | 0.01 | -1.20 | 0.24 | -0.03 | 0.01 | 0.01 |
| SA hide | SA self-blame rating bias | -0.01 | 0.01 | -0.73 | 0.47 | -0.02 | 0.01 | 0.00 |
|  | OA self-blame rating bias | 0.01 | 0.01 | 0.84 | 0.41 | -0.01 | 0.02 | 0.01 |
| SA distance from oneself | SA self-blame rating bias | 0.01 | 0.01 | -0.67 | 0.50 | -0.01 | 0.01 | 0.00 |
|  | OA self-blame rating bias | 0.00 | 0.00 | 0.50 | 0.62 | -0.01 | 0.01 | 0.00 |
| OA verbally attack friend | SA self-blame rating bias | -0.01 | 0.01 | -0.80 | 0.43 | -0.03 | 0.01 | 0.01 |
|  | OA self-blame rating bias | -0.05 | 0.01 | -6.95 | <.001***† | -0.06 | -0.04 | 0.26 |
| OA punish oneself | SA self-blame rating bias | 0.01 | 0.00 | 1.75 | 0.08 | 0.00 | 0.02 | 0.02 |
|  | OA self-blame rating bias | 0.02 | 0.00 | 5.13 | <.001***† | 0.01 | 0.02 | 0.16 |
| OA apologise | SA self-blame rating bias | 0.00 | 0.00 | 0.46 | 0.65 | -0.01 | 0.01 | 0.00 |
|  | OA self-blame rating bias | 0.01 | 0.00 | 4.56 | <0.001***† | 0.01 | 0.01 | 0.13 |
| OA hide | SA self-blame rating bias | 0.00 | 0.00 | 0.89 | 0.38 | -0.01 | 0.01 | 0.01 |
|  | OA self-blame rating bias | 0.01 | 0.00 | 2.17 | 0.03* | 0.00 | 0.01 | 0.03 |
| OA distance from self | SA self-blame rating bias | 0.01 | 0.00 | 1.63 | 0.11 | 0.00 | 0.01 | 0.02 |
|  | OA self-blame rating bias | 0.01 | 0.00 | 2.25 | 0.03* | 0.00 | 0.01 | 0.04 |
| OA distance from friend | SA self-blame rating bias | 0.02 | 0.01 | 1.52 | 0.13 | 0.00 | 0.04 | 0.02 |
|  | OA self-blame rating bias | 0.00 | 0.01 | -0.53 | 0.60 | -0.02 | 0.01 | 0.00 |

Self-blame rating biases were calculated by subtracting each participant's average other-blaming ratings from their average self-blaming ratings for each condition. The table included parameter estimates for post-hoc univariate analysis following significant multivariate effect, with self-blame bias for each agency condition as predictors and action tendencies as dependent variables. Positive regression coefficients (B) indicate a positive association between action tendencies and self-blaming biases. \*\*\* $p < .001$ , \*\* $p < .01$ , \* $p < .05$ at significance level of $\alpha = .05$ . † $p < .05$ after multiple comparison correction with Benjamini–Hochberg procedure. SA: self-agency; OA: other-agency.

### Group differences on action tendencies in the VR task

As predicted, a MANOVA showed a significant effect of group on the multivariate action tendency profile: F(10,127)=2.72, p=.005, Wilk’s Λ = .18, partial η2 = .18. Post-hoc univariate ANOVAs showed that compared with the control group, participants with depression had a higher proneness to feeling like punishing themselves and hiding irrespective of agency, a higher frequency of feeling like creating a distance from themselves in the other-agency and a lower frequency of feeling like apologising in the self-agency condition, as well as of verbally attacking one’s friend irrespective of agency (see Table 5 and Figure 3). After multiple comparison correction, depressed participants continued to show a significantly higher tendency to punish themselves in both agency conditions, as well as a higher tendency to hide and a lower tendency to verbally attack their friend in the other-agency condition compared with the control participants.

**Table 5.**
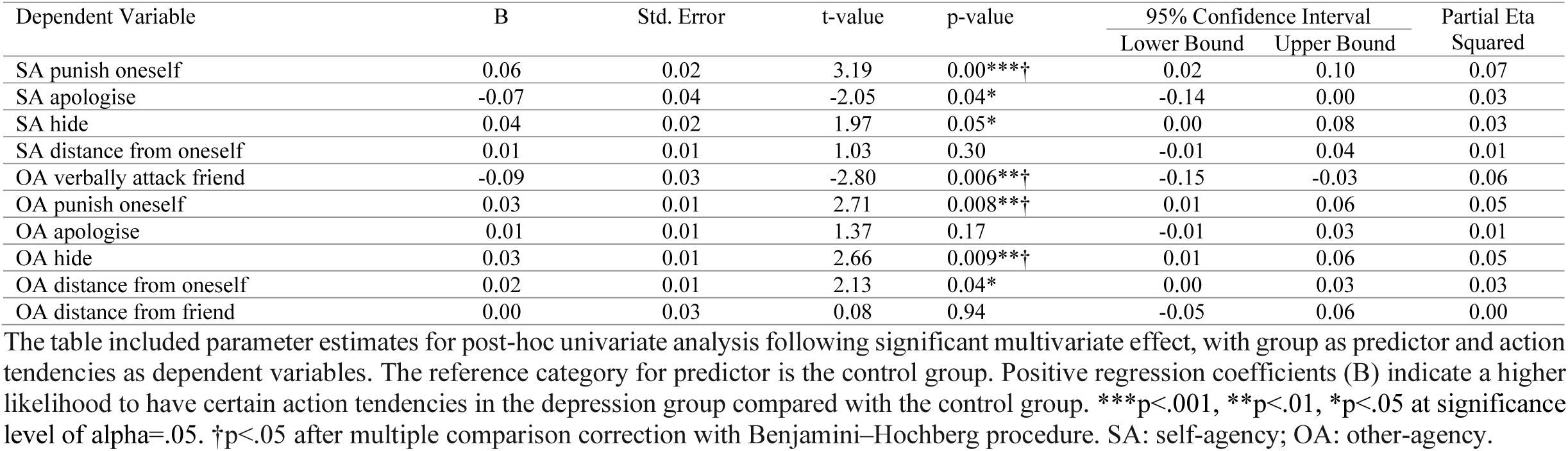
Parameter estimates for effects of group on action tendencies

**Figure 3.**
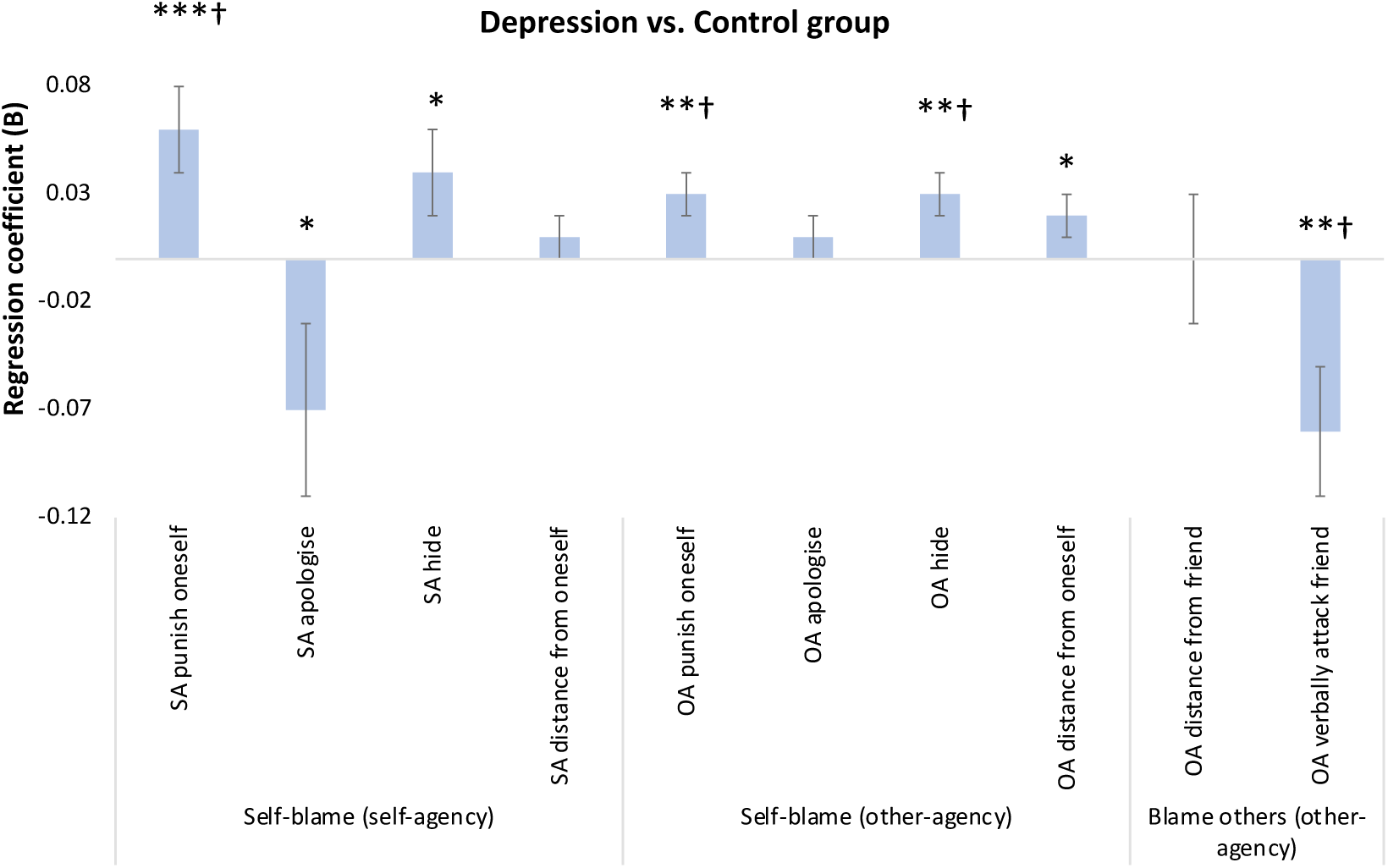
Regression coefficients for effects of group on action tendencies N=138 (depression group: 98; control group: 40); positive regression coefficients (B) indicate a higher likelihood to have certain action tendencies in the depression group compared with the control group. ***p<.001, **p<.01, *p<.05 at significance level of alpha=.05. †p<.05 after multiple comparison correction with Benjamini–Hochberg procedure. SA: self-agency; OA: other-agency.

### The role of feeling like punishing oneself in relation to self-harm and suicide attempts

Based on our finding that feeling like punishing oneself was the action tendency which most strongly distinguished the depression from the control group in both agency conditions, we investigated their relationship with reported history of self-harm or suicide attempts as recorded in our online baseline assessment. History of self-harm exhibited a significant multivariate association with feeling like punishing oneself irrespective of agency: F(2,91)=3.42, p=.037, Wilk’s Λ = .07, partial η2 = .07. Interestingly, this effect was not found for history of suicide attempts: F(2,91)=.46, p=.64, Wilk’s Λ = .01, partial η2 = .01. Post-hoc univariate ANOVAs showed that a previous history of self-harm was more specifically associated with an agency-incongruent (i.e. overgeneralised) feeling like punishing oneself when one’s friend had acted badly towards oneself (i.e. in the other-agency condition, see Table 6 and Figure 4).

**Table 6.** Differences in the tendency to punish oneself in relation to past self-harm and suicide attempts in the depression group

| Dependent Variable |  | B | Std. Error | t-value | p-value | 95% Confidence Interval |  | Partial Eta Squared |
| --- | --- | --- | --- | --- | --- | --- | --- | --- |
|  |  |  |  |  |  | Lower Bound | Upper Bound |  |
| SA punish self | self-harm | 0.03 | 0.03 | 1.11 | 0.27 | 0.02 | 0.08 | 0.01 |
|  | suicide attempt | 0.02 | 0.03 | 0.88 | 0.38 | -0.03 | 0.08 | 0.01 |
| OA punish self | self-harm | 0.04 | 0.02 | 2.58 | 0.01*† | -0.01 | 0.07 | 0.07 |
|  | suicide attempt | 0.01 | 0.02 | 0.59 | 0.55 | -0.02 | 0.04 | 0.00 |

The table included parameter estimates for post-hoc univariate analysis following significant multivariate effect, with self-harm and suicide attempt as predictors and punishing oneself for each agency as dependent variables. Reference categories for predictors are no self-harm and no suicide attempt. Positive regression coefficients (B) indicate a positive association between action tendencies and self-harm/suicide attempt. \* $p < .05$ at significance level of $\alpha = .05$ . † $p < .05$ after multiple comparison correction with Benjamini–Hochberg procedure. SA: self-agency; OA: other-agency.

**Figure 4.**
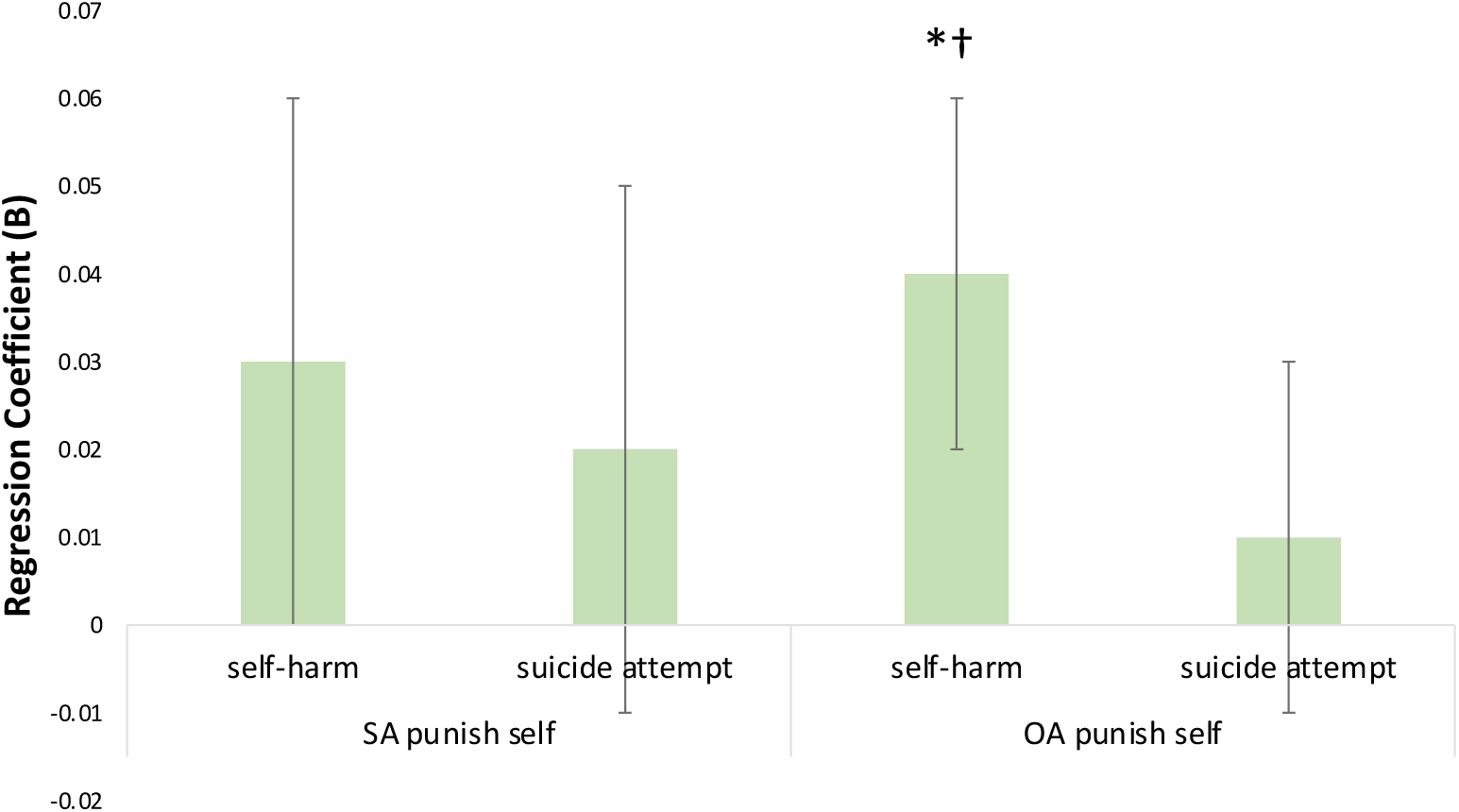
Tendencies to punish oneself in relation to past self-harm and suicide attempts in the depression group N=98; positive regression coefficients (B) indicate a positive association between action tendencies and self-harm/suicide attempt. ***p<.001, **p<.01, *p<.05 at significance level of alpha=.05. †p<.05 after multiple comparison correction with Benjamini–Hochberg procedure. SA: self-agency; OA: other-agency.

### Response time for action tendency choices in the VR task

The average time taken to choose action tendencies in each agency and group is shown in Supplementary Table 3. Interestingly, a repeated measure ANOVA revealed that all participants took a longer time to respond in the other-agency compared with self-agency condition as shown by a main effect of agency: F(1, 136=19.21), p=<.001. The discrepancy between conditions was greater in the depression group, as shown by an interaction between agency and group: F(1,136)=5.80, p=.02. There was no main effect of group: F(1,136)=.81, p=.37.

## Discussion

In the present study, we developed a novel VR task to assess blame-related action tendencies. The first aim of the study was to validate the novel VR task as a self-assessment measure of blame-related action tendencies and self-blaming biases in people with current depression by comparing it against a shortened, optimised version of our previously reported text-based version (Duan et al., 2022). Our results showed that most action tendencies and self-blaming biases in the VR task were significantly correlated with those in the text-based task, which suggested good concurrent validity of the novel VR task. This allowed us to probe our second hypothesis that people with current depression exhibit higher levels of maladaptive blame-related action tendencies, including feeling like hiding, creating a distance from oneself and self-punishing. Our results confirmed this hypothesis by showing that the depression group exhibited a higher proneness to all hypothesized maladaptive blame-related compared with the control group at a multivariate level and was most pronounced for feeling like punishing oneself and hiding. Although the group difference for creating a distance from oneself did not survive multiple comparison correction, the trend was still consistent with our prediction and also in line with previous findings that people with depression had a tendency to imagine things from a third-person perspective (Holmes et al., 2016). Furthermore, people with depression showed reduced adaptive feelings of verbally attacking one’s friend when they were described to have acted badly towards them, which is consistent with previous findings of reduced anger towards others in remitted depression (Zahn et al., 2015). Our third aim was to investigate the hypothesis that maladaptive action tendencies, specifically feeling like punishing oneself, are associated with a higher risk of self-harm and suicide attempt based on self-reported history. This hypothesis was partially supported by our findings that participants with higher likelihood of punishing oneself had higher rate of self-harm, but not of suicide attempts.

Our finding that the contextualised VR-scenarios concord with abstract single sentence descriptions in the text-based task provides construct validity for the measures of higher maladaptive action tendencies including feeling like hiding and self-punishing in depression. Importantly, this is the first demonstration of maladaptive self-blame-related action tendencies in people with current depression who are also likely to exhibit a much higher level of co-morbid conditions such as anxiety disorders, thus providing crucial evidence for the generalisability to non-remitting forms of depression. Although the link between depression and maladaptive action tendencies requires further investigation, it might reflect depressogenic coping styles. As action tendencies are implicit motivational states before an action is taken (Haidt, 2003), they are highly likely to play a role in subsequent behaviour (Roseman et al., 1994). As a result, people with a tendency to hide and self-punish might adopt maladaptive actions as coping methods, such as avoidance and self-harm. These coping methods may relieve tension in the short term, but reduce the likelihood of problem-solving in the long term, which ultimately contribute to depressogenic schemata (Beck & Haigh, 2014).

Our analysis of response times intriguingly showed a longer response time in the other-agency compared with the self-agency condition which was significantly more pronounced in the depression group. This could reflect a higher level of ambivalence about their choices in the other-agency condition. The other-agency condition was also where people with depression showed the most distinctive profile of agency-incongruent self-blame-related action tendencies, particularly feeling like punishing oneself. The finding that a higher likelihood of punishing oneself was associated with a higher likelihood of a history of self-harm, but not of suicide attempts, is consistent with previous findings that self-harm in depression was related to punishment-related behaviours (Stanicke, 2021). Given the high prevalence and the detrimental effect of self-harm in people with depression, especially adolescents (Stallard et al., 2013), our finding revealed a novel target for treatment planning and the development of prevention strategies of self-harm in depression. As feeling like self-punishing was not associated with suicide attempts, this points to self-harm as a more specific clinical feature with a distinct motivational underpinning. Although studies found that a history of self-harm increased the likelihood of suicide attempt, it was not specifically linked with thoughts of dying (Mitchell & Dennis, 2006). Self-harm, unlike suicidality, is also less likely linked to hopelessness, shown to be of particular relevance for suicidality (Abramson et al., 1989).

### Limitations

On a more cautionary note, our study was limited by not including a diagnostic interview and so we were unable to establish a formal diagnosis of current major depressive disorder (MDD). It is, however, highly likely that our depression group consists almost exclusively of people who would fulfil MDD criteria, given that: 1) we only included participants who were deemed to require pharmacological treatment of their depression by their GP, and used a highly conservative PHQ-9 score cut-off (≥ 15), which has a specificity of .96 for MDD (Manea et al., 2012); 2) used a rigorous way of excluding people with a bipolar history, and 3) used validated scales for excluding substance and alcohol use disorders. A further limitation was the cross-sectional design of the study, which did not allow us to examine the causal relationships between maladaptive action tendencies and depression. Longitudinal studies are needed to examine state and trait-related aspects of maladaptive action tendencies. The fact that we found no association of symptom severity with action tendencies suggests that these reflect vulnerability traits rather than states. Due to recruiting a pragmatic sample of people who have not benefitted from antidepressant treatments, we were unable to rule out the effect of medication, although we did not find an influence of medication status on action tendencies.

### Conclusions

Taken together, our self-administered VR task of blame-related biases showed good construct validity and excellent suitability for remote use in depression. As shown by the task, feeling like hiding and self-punishing were distinctive for participants with depression compared with those without depression, consistent with our previous findings based on a text-based assessment. In addition, feeling like punishing oneself was specifically associated with a history of self-harm, but not suicide attempts in people with depression. Our finding unveils novel cognitive markers, neurocognitive prevention and treatment targets, as well as provides the first step in validating the task as a measure of self-blaming biases in depression.

## Data Availability

All data produced in the present study are available upon reasonable request to the authors

## Author Contributions

SD and RZ developed the study concept. All authors contributed significantly to the study design. Data was collected by SD and DF. Testing and analysis were performed by SD under the supervision of RZ. SD drafted the manuscript, and LV, DF, JM and RZ provided critical revisions. All authors approved the final version of the manuscript for submission.

## Acknowledgement

We greatly thank Jerome Di Pietro for the software development of the VR assessment in this study.

