## Supplementary materials for "Remote virtual reality assessment elucidates self-blame-related action tendencies in depression"

[3] *South London and Maudsley NHS Trust*

[4] *Ku Leuven, Belgium*

[5]*Cognitive and Behavioral Neuroscience Unit, D'Or Institute for Research and Education (IDOR), 22280-080 - Rio de Janeiro, RJ, Brazil*

[6] *Scients Institute, USA*

*Corresponding author

Dr Roland Zahn (see address above)

**Supplementary Figure 1|** A flow chart of participant recruitment

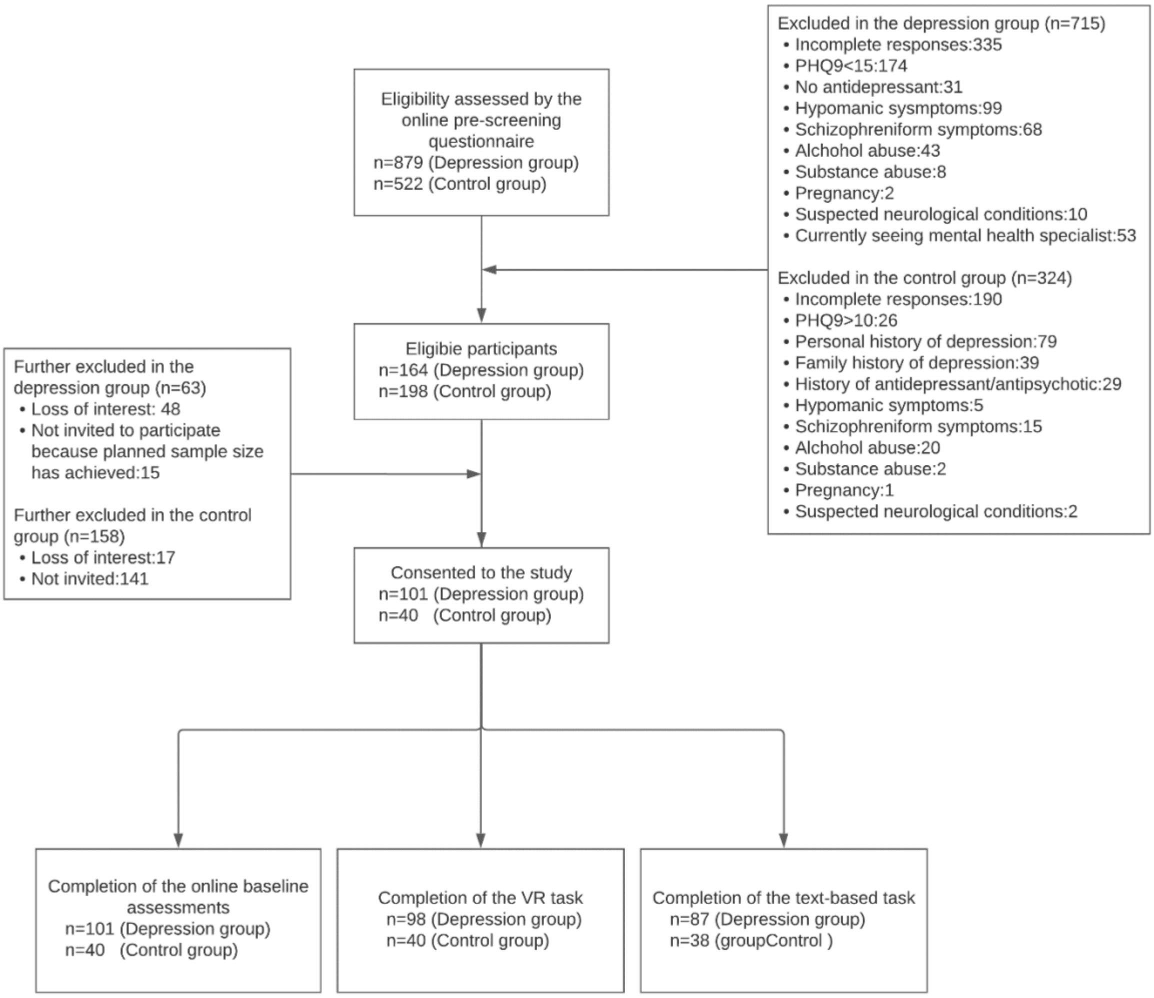

**Supplementary Table 1|** Scenarios used in the VR task

| No. | Self-agency condition | Other-agency condition |
|---|---|---|
| 1. | You drove your friend's car, caused an accident, and damaged it. | Your friend drove your car, caused an accident, and damaged it. |
| 2. | At your friend's party, you stained their carpet. | At your party, your friend stained your carpet. |
| 3. | You spoke negatively about your friend to their boss. | Your friend spoke negatively about you to your boss. |
| 4. | At your friend's house, you cheated during a game. | At your house, your friend cheated during a game. |
| 5. | When babysitting for your friend, you shouted at their child. | When babysitting for you, your friend shouted at your child. |
| 6. | Your friend lent you money, and you did not pay them back. | You lent your friend money, and they did not pay you back. |
| 7. | Your friend caught you in a lie | You caught your friend in a lie |
| 8. | Whilst your friend was on holiday, you kissed their partner. | Whilst you were away on holiday, your friend kissed your partner. |
| 9. | When with other friends, you exposed your friend's secret. | When with other friends, your friend exposed your secret. |
| 10. | You arrived very late to a lunch with your friend and ruined some of the fun. | Your friend arrived very late to a lunch with you and ruined some of the fun. |
| 11. | You have gossiped about your friend. | Your friend has gossiped about you |
| 12. | To avoid seeing your friend, you lied about your plans. | To avoid seeing you, your friend lied about their plans. |
| 13. | During a disagreement, you swore at your friend. | During a disagreement, your friend swore at you. |
| 14. | When your friend rang the doorbell, you pretended not to be at home. | When you rang the doorbell, your friend pretended not to be at home. |
| 15. | When your friend needed some help, you did not lend a hand. | When you needed some help, your friend did not lend a hand. |

**Supplementary Table 2|** Pearson correlations between action tendencies and symptoms in the depression group

| Group | | Self-agency | | | | Other-agency | | | | | |
|---|---|---|---|---|---|---|---|---|---|---|---|
| | | Punish self | Apologize | Hide | Distance from self | Verbally attack friend | Punish self | Apologize | Hide | Distance from self | Distance from friend |
| QIDS-SR-16 | r | 0.03 | 0.03 | 0.01 | -0.14 | 0.01 | 0.08 | -0.10 | -0.04 | -0.15 | 0.12 |
| | p | 0.76 | 0.77 | 1.00 | 0.17 | 0.92 | 0.44 | 0.33 | 0.70 | 0.16 | 0.25 |
| GAD-7 | r | 0.02 | -0.05 | 0.06 | -0.10 | 0.12 | -0.04 | -0.09 | -0.15 | 0.01 | 0.02 |
| | p | 0.83 | 0.60 | 0.5 | 0.33 | 0.26 | 0.71 | 0.39 | 0.14 | 0.92 | 0.89 |

n=98 for the depression group and n=40 for the control group. QIDS-SR-16: Quick Inventory of Depressive Symptomatology-Self-Reported-16; GAD-7: generalised anxiety disorders-7 scale, r= Pearson’s correlation coefficient, p=uncorrected 2-sided p-values. No correlation was significant at an uncorrected p=.05 2-sided.

**Supplementary Table 3|** Response times (seconds) for action tendencies in each agency condition and group

| Agency | Group | Mean | Std. Deviation |
|---|---|---|---|
| Self-agency | Control | 7.14 | 3.19 |
| | Depression | 7.04 | 4.17 |
| | Both | 7.07 | 3.90 |
| Other-agency | Control | 7.86 | 2.94 |
| | Depression | 9.50 | 6.68 |
| | Both | 9.02 | 5.88 |

n=98 for the depression group and n=40 for the control group.
